# The effect of BCG vaccination on COVID-19 examined by a statistical approach: no positive results from the Diamond Princess and cross-national differences previously reported by world-wide comparisons are flawed in several ways

**DOI:** 10.1101/2020.04.17.20068601

**Authors:** Masakazu Asahara

**Affiliations:** Division of Liberal Arts and Sciences, Aichi Gakuin University, Nisshin, Aichi 470-0195, Japan

## Abstract

Recently, the controversial hypothesis that past BCG (Bacillus Calmette–Guérin) vaccination reduces infection or severity of COVID-19 has been proposed. The present study examined this hypothesis using statistical approaches based on the public data. Three approaches were utilized: 1) comparing the infection and mortality ratio of people on the cruise ship Diamond Princess, 2) comparing the number of mortalities among nations, and 3) comparing the maximum daily increase rate of total mortalities among nations. The result of 1) showed that there is no significant difference in infection per person onboard or mortality-infection between Japanese citizens vs. US citizens and BCG obligatory nations vs. non-BCG obligatory nations on the Diamond Princess. The result of 2) showed that the number of mortalities among nations is similar to the previous studies, but this analysis also considered the timing of COVID-19 arrival in each nation. After correcting for arrival time, previously reported effect of BCG vaccination on decreasing total mortality disappeared. This is because nations that lack BCG vaccination are concentrated in Western Europe, which is near an epicenter of COVID-19. Therefore some previous reports are now considered to be affected by this artifact; the result may have been flawed by dispersal from an epicenter. However, some results showed weakly significant differences in the number of deaths at a particular time among BCG obligatory and non-BCG nations (especially the use of Japanese BCG strain Tokyo 172). However, these results are affected by the results of three countries and the effect of BCG vaccination remains inconclusive. The result of 3) showed that the maximum daily increasing rate in death among nations showed no significant difference among BCG vaccination policies. In the present study, although some results showed statistically significant differences among BCG vaccination policies, they may be affected by the impact of various other factors, such as national infection-control policies, social distancing, behavioral changes of people, possible previous local epidemics of closely related viruses, or inter-population differences in *ACE2* or other genetic polymorphism. Further research is needed to better understand the underlying cause of the observed differences in infection and mortality of the disease among nations. Nevertheless, our results show that the effect of past BCG vaccination, if any, can be masked by many other factors. Therefore, the possible effect might be relatively small. In fact, in Japan, where almost all citizens have been vaccinated, COVID-19 cases are constantly increasing. Given the importance of people’s behavior in preventing viral propagation, the spread of optimism triggered by this hypothesis would be harmful to BCG vaccination nations.

## Introduction

In April 2020, the global pandemic of coronavirus disease-2019 (COVID-19) caused by severe acute respiratory coronavirus 2 (SARS-CoV-2) is a serious problem all over the world. Recently, several authors have suggested the hypothesis that past BCG (Bacillus Calmette-Guérinvaccination) vaccination reduces the morbidity and mortality of COVID-19. Several authors reported the correlation between national BCG vaccination policy and infection or mortality rate (or its increasing rate) in those nations (Akiyama and Ishida 2020; Berg et al. 2020; Dolgikh 2020; Goswami et al. 2020; Miller et al. 2020; Sala and Miyakawa 2020; Shet et al. 2020), and some have argued that the hypothesis is supported. However, several authors failed to account for how infectious diseases spread from one place to another. That is, several authors simply compared the total number of deaths in each nation without considering the timing disease arrival in each nation (e.g. Miller et al. 2020; Sala and Miyakawa 2020), suggesting that past BCG vaccination changes the number of infection and death by double or triple digits (e.g. they simply compared 16523 deaths until April 6 in Italy which BCG- vs. 47 deaths until April 6 in Russia which BCG+, where the timing of the pandemic enter to the nation is different). This shortcoming would produce critically important misunderstandings of the effect of BCG vaccination on COVID-19. In fact, mandatory BCG vaccination is discontinued in Western Europe, which is near a COVID-19 epicenter. This should affect the conclusions of previous studies. Therefore, the present study attempted to examine the effect of BCG vaccination while accounting for these biases.

Three approaches were used in the present study. The first approach is a test in the cruise ship Diamond Princess. In this ship, most patients were infected before they were aware that the disease was spreading in the ship. Therefore, the cultural effect of countries (such as wearing masks) or national policy (such as the “cluster buster” policy in Japan) can be excluded. The strain of virus on the ship is the same, and the spread occurred simultaneously on board (Sekizuka et al. 2020). The second approach is a similar comparison as in many previous studies (e.g. Miller et al. 2020; Sala and Miyakawa 2020). That is, the number of mortalities in each nation was compared, but with consideration for the timing of disease arrival. The third approach is also similar to a previous study (Akiyama and Ishida 2020). That is, the rate of increase of the number of mortalities in each nation was compared.

## Materials and Methods

Data was obtained from previous publications or public databases. The number of cases, deaths, and passengers’ nationality on the Diamond princess was obtained from public data (some districts within the nation were treated separately) (Ministry of Health, Labour and Welfare, Japan, 2020a; b) and a previous report (Moriarty et al. 2020). BCG vaccination policies were obtained from The BCG World Atlas (Zwerling et al. 2011; http://www.bcgatlas.org/). The nationalities of dead passengers were obtained from each news report.

Data on cases and mortality from each nation was obtained from a public database (some districts within the nation were treated separately) (Dong et al. 2020; Johns Hopkins University). The number of international tourist' arrivals in 2017 was obtained from a previously published report (World Tourism Organization 2019). Some data (BCG vaccination policy, population, and life expectancy) were obtained from the supplementary material of a previous study (Sala and Miyakawa 2020). Some BCG strain information was obtained from other literature (Akiyama and Ishida 2020; Joung and Ryoo, 2013).

In the analysis of the Diamond Princess, the effects of BCG policy and nationality were examined using the Fisher’s exact test. If the BCG vaccination changes the infection or mortality rate by two or three digits as previous studies suggested, a power analysis indicated that the sample size for the analysis of infection rate may be sufficient for 90% power on the analysis (e.g., n=62 is sufficient to detect a difference between the incidence rates of 0.2 and 0.02 in each group). With respect to the mortality rate, the sample size may meet 50 to 70% of the power on the analysis (e.g., 70%: n=124 is sufficient for 70% power to detect a difference between the incidence rates of 0.05 and 0.0005; 60%: n=99, 0.05 and 0.0005; 50%: n=394, 0.01 and 0.0001; n=78, 0.05 and 0.0005, respectively).

To compare the cases and deaths among nations, the timing of the fifth death was used to align the timing of disease entry to the nation. Although Shet et al. (2020) used the timing of the 100^th^ case, this alignment is considered better because the specific number of the case depends on the national policy of examination. In addition, the present study focused on the total number of deaths in each nation, rather than death per population, because the pandemic was severe in particular cities or particular areas. In other words, it does not make sense to compensate for the spread of infection in Hubei province with the population of Beijing and Shanghai. This relationship is the same in Moscow and Siberia, and the same in Tokyo and Iwate. Moreover, national political decisions to prevent disease spread, which is one of the most important factors of disease spread, should be affected by the total number of victims. Therefore, aligning to the total national population is irrelevant. Moreover, almost all people are still considered susceptible to the disease (even in small nations such as San Marino). In some nations, such as China, the increase in the number of deaths was stopped or flattened. Therefore, cumulative deaths 30 days after the date fifth death (this treatment is based on Akiyama and Ishida 2020) was used. The general linear model was used to test the effect of past BCG vaccination, the timing of pandemic entry to the nation, and region (e.g., East Asia, Europe, etc.). Numbers of death were log-transformed. The region may serve as rough indexes of social custom, different ratios of genetic polymorphisms, or the distribution of wildlife. For some results, nations using the Japanese BCG strain (Tokyo 172) were separated from other BCG strains because some reports have suggested the Japanese strain is more effective (Akiyama and Ishida 2020).

To compare the increasing rate of death in each nation, daily increases in the number of total deaths were calculated. The present study used two weeks average of the daily rate: Average rates were calculated every two weeks and the highest value was used as the maximum increase rate per day. Comparisons were performed using nations where the total number of deaths was over 25. The effect of BCG policy and region was examined using the ANOVA-Tukey test. A posthoc power analysis indicated that if the between-group variance was over 0.0075 (Japanese strain unseparated) and 0.021 (Japanese strain separated), the sample size is sufficient for 90% power on the analysis. These variances are comparable to within-group variances. The data of Serbia may not be completed, it was excluded from this analysis. In this study, all statistical analyses were performed by Minitab 18 (Minitab Inc, USA) except for the power analyses which were performed by R (R Core Team, Vienna, Austria). The data and calculations in the spreadsheet are shown in the supplementary data.

## Results and Discussion

### The Diamond Princess shows no positive perspective for a personal-level effect of BCG vaccination

The results of the Diamond Princess are shown in Table 1 and 2. The case-fatality ratio was not significantly different between people from BCG mandatory or not mandatory nations even when the analysis was limited to the elderly (Table 1). The infection and mortality ratio were not significantly different between Japanese citizens (BCG+) and US citizens (BCG-) (Table 2). The mortality rate was not significantly different between passengers from several countries (Table 2).

**Table 1.** Case fatality ratio between the nationality of different BCG vaccination policies in the Diamond Princess (April 27, 2020). Both past and current implementing of vaccination are categorized to + Current implementing of vaccination is categorized to + *There are still 6 people in the hospital whose nationality is undisclosed.

| Both past and current implementing of vaccination are categorized to + |  |  |  |
| --- | --- | --- | --- |
| BCG | Cases | Deaths | Mortality rate (%) |
| + | 493 | 13 | 2.64 |
| – | 141 | 1 | 0.71 |

| BCG | Cases | Deaths | Mortality rate (%) |
| --- | --- | --- | --- |
| + | 422 | 11 | 2.61 |
| – | 212 | 3 | 1.42 |

| Both past and current implementing of vaccination are categorized to + |  |  |  |
| --- | --- | --- | --- |
| BCG | Cases | Deaths | Mortality rate (%) |
| + | 350 | 13 | 3.71 |
| – | 126 | 1 | 0.79 |

| BCG | Cases | Deaths | Mortality rate (%) |
| --- | --- | --- | --- |
| + | 301 | 11 | 3.65 |
| – | 175 | 3 | 1.71 |

| Both past and current implementing of vaccination are categorized to + |  |  |  |
| --- | --- | --- | --- |
| BCG | Cases | Deaths | Mortality rate (%) |
| + | 227 | 13 | 5.73 |
| – | 71 | 1 | 1.41 |

| BCG | Cases | Deaths | Mortality rate (%) |
| --- | --- | --- | --- |
| + | 198 | 11 | 5.56 |
| – | 100 | 3 | 3.00 |

No significant difference p=0.398 (Fisher's exact test)
\*There are still 8 people in hospital whose nationality is undisclosed

**Table 2.** Infection and mortality ratio between citizens of Japan and other countries in the Diamond Princess (April 27, 2020)

| Nationality | Number on board | Cases | Infection rate (%) |
| --- | --- | --- | --- |
| Japan (BCG +) | 1341 | 270 | 20.13 |
| USA (BCG -) | 428 | 107 | 25.00 |

| Nationality | Number on board | Deaths | Mortality rate (%) |
| --- | --- | --- | --- |
| Japan (BCG +) | 1341 | 9 | 0.67 |
| USA (BCG -) | 428 | 0 | 0.00 |

| Nationality | Number on board | Deaths | Mortality rate (%) |
| --- | --- | --- | --- |
| Japan (BCG +) | 1281 | 9 | 0.70 |
| Hong Kong (BCG +) | 260 | 2 | 0.77 |
| USA (BCG -) | 416 | 0 | 0.00 |
| Canada (BCG -) | 251 | 1 | 0.40 |
| Australia (BCG - / past+) | 223 | 1 | 0.45 |
| UK (BCG - / past+) | 57 | 1 | 1.75 |

No significant difference Japan + Hong Kong vs Others; Others vs USA + Canada: p > 0.05 (Fisher's exact test)
\*1 There are still 8 people in hospital whose nationality is undisclosed
\*2 US data was obtained from Moriarty et al. (2020)
\*3 Passengers data was obtained from Moriarty et al. (2020)

To discuss the result, Japanese citizens under age 98 should have received BCG vaccination; in 1951 all infants and citizens under 29 were inoculated BCG vaccine if their tuberculin test was negative (Ministry of Health, Labour and Welfare, Japan, 2016). According to the estimated rate of tuberculosis infection, 12.0% of five year old children and 24.6% of ten year old children were infected with tuberculosis in 1950 (Omori 2009). Therefore, 80-90% of 70-79 years old people were considered to be inoculated with the BCG vaccine in Japan. While most Japanese victims on the Diamond Princess were over 70 years old (except for two victims whose age has not been announced), most had been inoculated with the BCG vaccination. However, the infection rate and mortality rate were not different significantly from that of other people onboard from BCG negative nations.

It should be noted that there was 1045 staff among the 3711 people on the Diamond Princess (National Institute of Infectious Diseases, 2020). Staff may be younger than passengers, and the mortality rate should be relatively low. However, when the analyses focused on older people or passengers only, the conclusion was not changed (Table 1 and 2). In addition, at that time, the Diamond Princess traveled from Hong Kong to Japan. Therefore, East Asian people (from BCG+ countries) could have easily boarded the cruise, even if they had pre-existing diseases. However, passengers from Western Europe, the United States, and Australia may have been restricted to individuals without pre-existing diseases due to the long flight required to reach the cruise. In addition, 8 people remain in the hospital as of 2020 April 27 (Ministry of Health, Labour and Welfare, Japan, 2020a). These factors may affect the result. However, it should be emphasized that the Diamond Princess presented no positive support for the hypothesis that past BCG vaccination reduces the infection and mortality of COVID-19.

### Cross-national comparison showed that previous reports were flawed due to the timing of spread, yet there is still a weakly significant result in particular comparisons

The bivariate plots show the results of the cross-national comparison (Table 3 and 4; Fig. 1, 2, 3, 4, and 5). The result indicated that the number of deaths is seriously affected by when the disease first spread to the nation (i.e. date of the fifth death) (Table 3 and 4). In addition, it is also significantly affected by the region which is used as an index of social custom, different ratios of genetic polymorphisms, or the distribution of wildlife. For the total number of deaths, when the period after pandemic entry was fixed, the BCG effect disappeared (Analysis A and C: Table 3). In contrast, when analyzing the Japanese BCG strain separately, BCG policy was not significant in analysis B, but was significant in analysis D (Table 3). This result may be due to three nations (Japan, Iraq, and South Korea) which showed relatively lower mortality and use the Japanese BCG strain. Therefore, unique characteristic effects of those nations may affect the result. For example, public security problems in Iraq might force people to stay home. The South Korean government examined many patients and succeeded in controlling the pandemic. The Japanese government adopted a unique strategy of “cluster buster” based on the previous result that the “super-spreader” (an infected person who spreads the virus to many other people) is restricted in SARS-CoV-2 (Nishiura et al. 2020). In addition, governmental requests for social distancing may work in Japan (on April 11, the number of train passengers in Shibuya station, Tokyo reduced 98% from one year ago, except for those using commuter’s ticket: TBS News 2020). The same metrics showed 66-89% of passenger reduction at other major stations in Tokyo that weekend (Cavinet Secretariat, Japan 2020). It should be noted that South Korea had been using the Danish BCG strain, but later changed to the Japanese strain Tokyo 172 (Akiyama and Ishida 2020). Among other nations examined, China and the Philippines, which are BCG+, showed relatively fewer deaths (Fig. 1 and 2). These nations are known to choose strong policies to prevent transfer among cities. Australia, which is a BCG-nation, also showed a similar pattern to Japan, Iraq, and South Korea (Fig. 1 and 2) although the cause of this is not clear. The pattern of San Marino may be explained by its smaller population. All nations possess particular factors that affect their results.

**Table 3.** The result of general linear model testing for the effect of BCG vaccination and other factors using all available nations around the world (Fig. 1 and 2)

| Log Total Death = BCG polocy + Date 5th death + Region |  |  |  |  |
| --- | --- | --- | --- | --- |
|  | Adj SS | Adj MS | F | P |
| BCG policy | 0.4857 | 0.2429 | 1.730 | 0.184 |
| Date 5th death | 29.1332 | 29.1332 | 207.920 | 0.000 |
| Region | 8.1498 | 1.0187 | 7.270 | 0.000 |
| R<sup>2</sup>=0.8367 |  |  |  |  |

Analysis C
| Log Period Death = BCG polocy + Days from 5th death + Region |  |  |  |  |
| --- | --- | --- | --- | --- |
|  | Adj SS | Adj MS | F | P |
| BCG policy | 0.0821 | 0.0411 | 0.280 | 0.760 |
| Days from 5th death | 25.005 | 25.005 | 168.160 | 0.000 |
| Region | 3.6858 | 0.4607 | 3.100 | 0.005 |
| R<sup>2</sup>=0.8080 |  |  |  |  |

\*Period Death: Total deaths or sum of deaths 30th days from 5th death

Analysis B
| Log Total Death = BCG polocy (J strain separated) + Date 5th death + Region |  |  |  |  |
| --- | --- | --- | --- | --- |
|  | Adj SS | Adj MS | F | P |
| BCG policy (Japanese strain separated) | 0.4895 | 0.1632 | 1.150 | 0.336 |
| Date 5th death | 29.006 | 29.006 | 204.210 | 0.000 |
| Region | 7.0906 | 0.8863 | 6.240 | 0.000 |
| R<sup>2</sup>=0.8091 |  |  |  |  |

Analysis D
| Log Period Death = BCG polocy (J strain separated) + Days from 5th death + Region |  |  |  |  |
| --- | --- | --- | --- | --- |
|  | Adj SS | Adj MS | F | P |
| BCG policy (Japanese strain separated) | 1.5887 | 0.5296 | 4.110 | 0.010 |
| Days from 5th death | 26.353 | 26.353 | 204.720 | 0.000 |
| Region | 2.4454 | 0.3057 | 2.370 | 0.026 |
| R<sup>2</sup>=0.8362 |  |  |  |  |

**Table 4.** The result of general linear model testing for the effect of BCG vaccination and other factors using European nations (Fig. 4)

|  | Adj SS | Adj MS | F | P |
| --- | --- | --- | --- | --- |
| BCG policy | 0.7688 | 0.3844 | 2.850 | 0.072 |
| Date 5th death | 17.3209 | 17.3209 | 128.450 | 0.000 |
| R<sup>2</sup>=0.8582 |  |  |  |  |

Analysis F Log Period Death = BCG polocy + Days from 5th death
|  | Adj SS | Adj MS | F | P |
| --- | --- | --- | --- | --- |
| BCG policy | 0.36 | 0.18 | 1.270 | 0.295 |
| Days from 5th death | 16.118 | 16.118 | 113.400 | 0.000 |
| R<sup>2</sup>=0.8427 |  |  |  |  |

\*Period Death: Total deaths or sum of deaths 30th days from 5th death

**Fig. 1.**
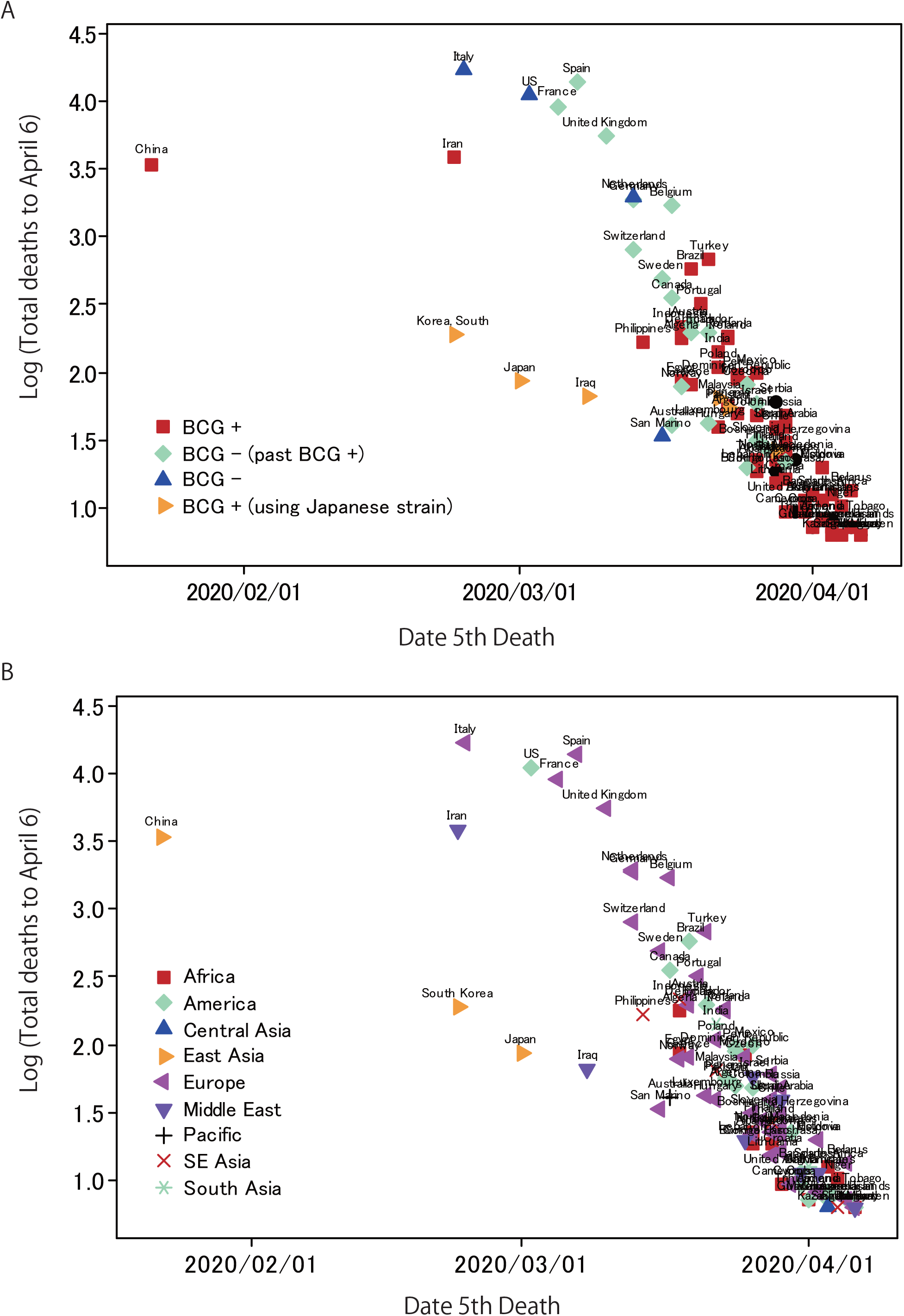
Bivariate plots of the Date of fifth Death (as an index of the timing of pandemic entry into the nation) vs. Log (Total deaths up to April 6). Most nations showed exponential increases in the number. However, some nations plotted lower than other nations. It should be noted that BCG – countries are mainly located in Europe, there is an epicenter of the pandemic.

**Fig. 2.**
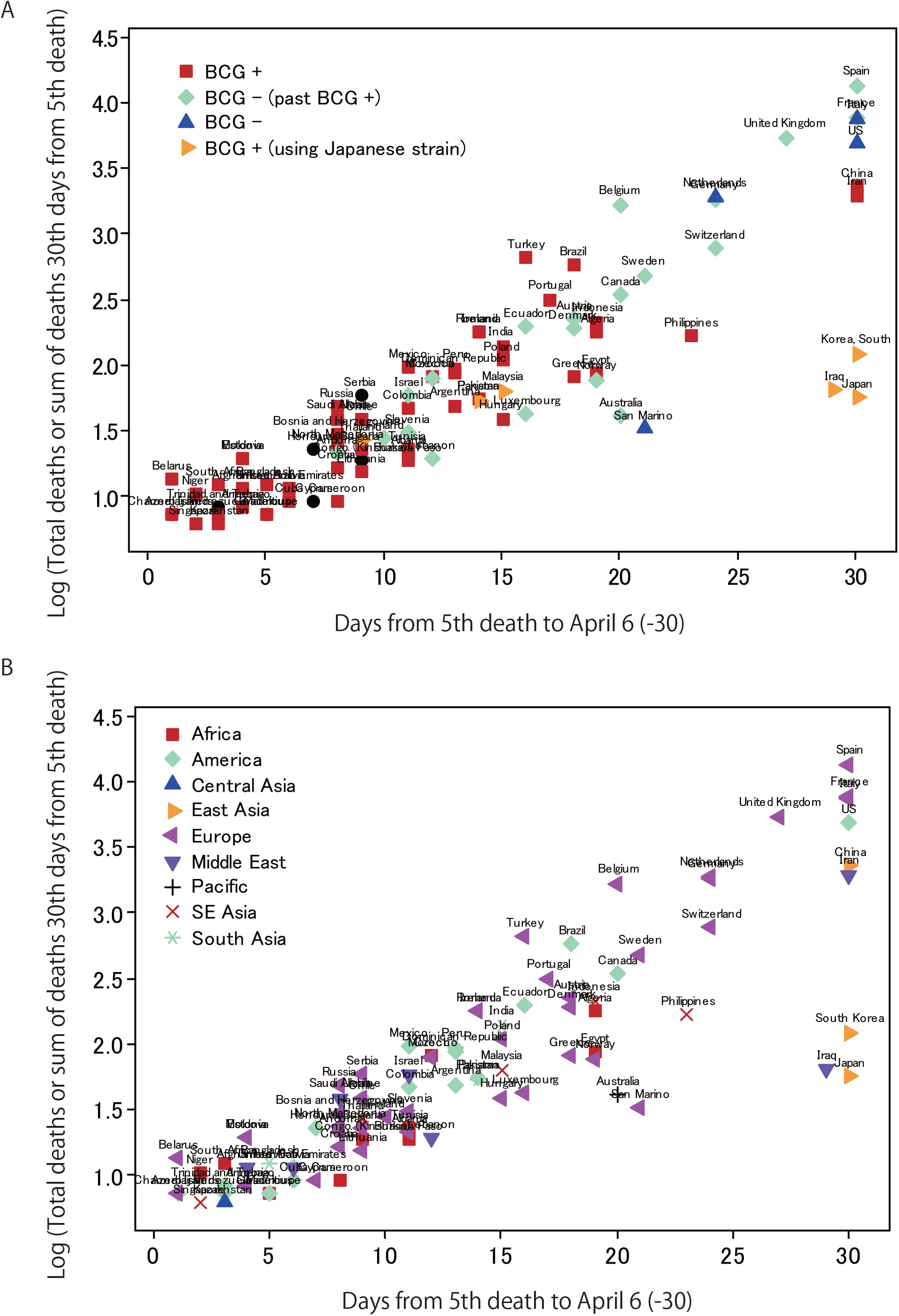
Bivariate plots of Days from fifth Death to April 6 or 30 (as an index of the time from the entry of the pandemic to the nation/the time until the number of deaths has flattened) vs. Log (Total deaths or sum of deaths at 30 days after the fifth death, i.e. the number of deaths during the time X axis). Most nations showed exponential growth in the number. However, some nations plotted lower than the other nations.

**Fig. 3.**
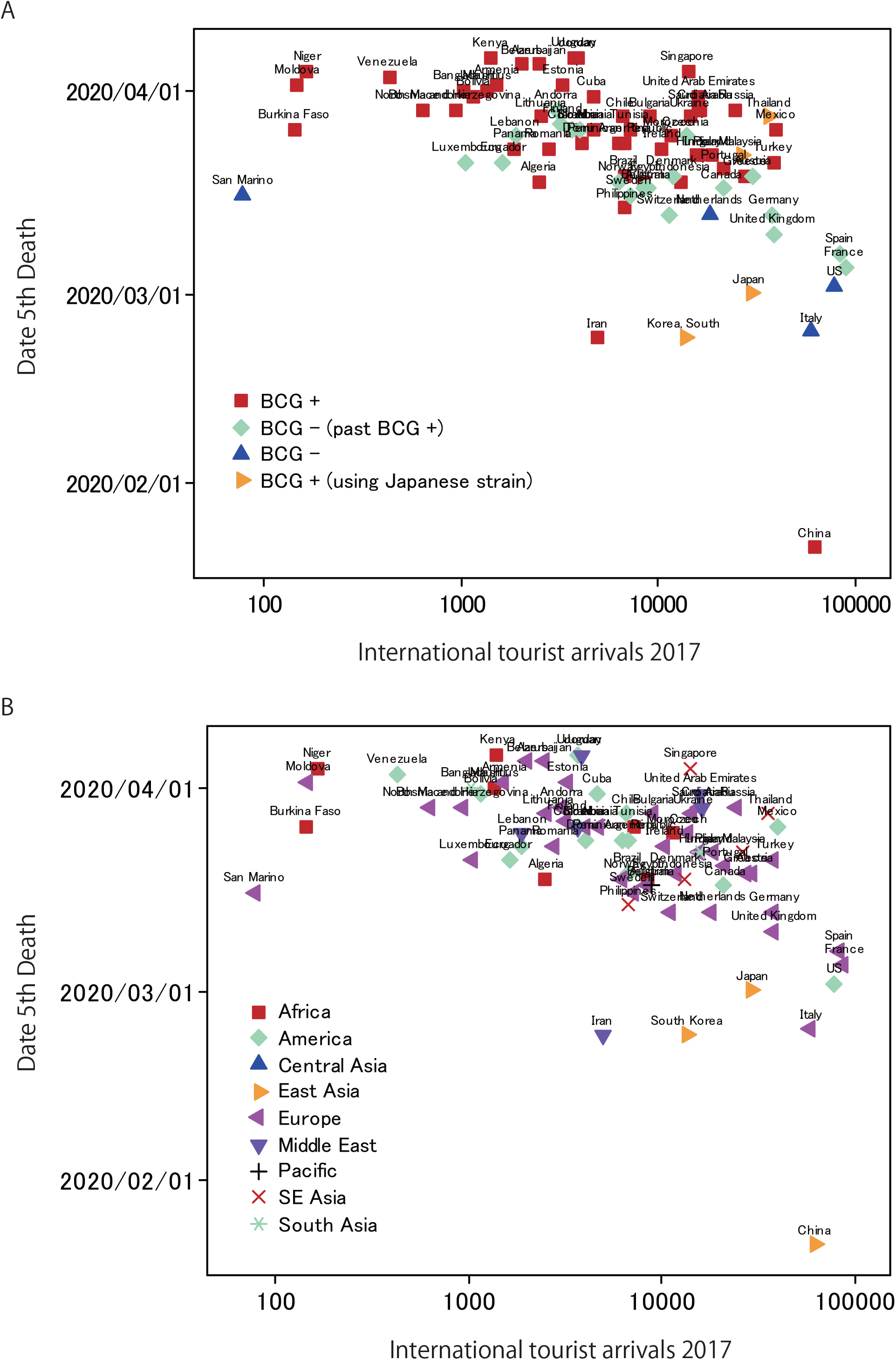
Bivariate plots of international tourist arrivals in 2017 vs. Date fifth Death. Pandemic propagation may be affected by tourist arrivals.

**Fig. 4.**
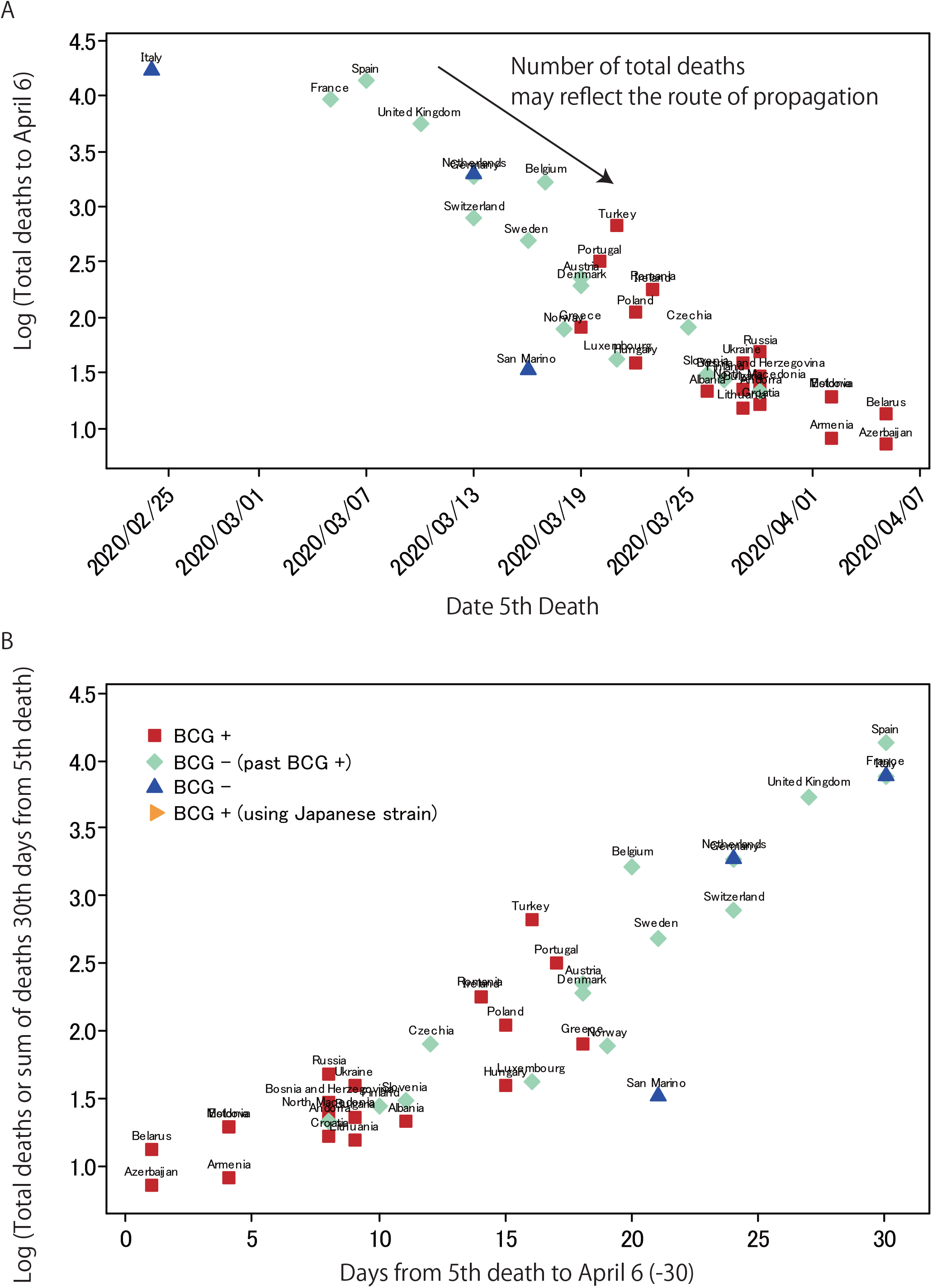
Bivariate plots using only European nations. The plots clearly show that high number of deaths are affected by when the pandemic enter the nation. The pandemic explosion occurred in Western Europe where BCG – nations are located, and then spread to the peripheral of Europe where BCG + nations are located.

**Fig. 5.**
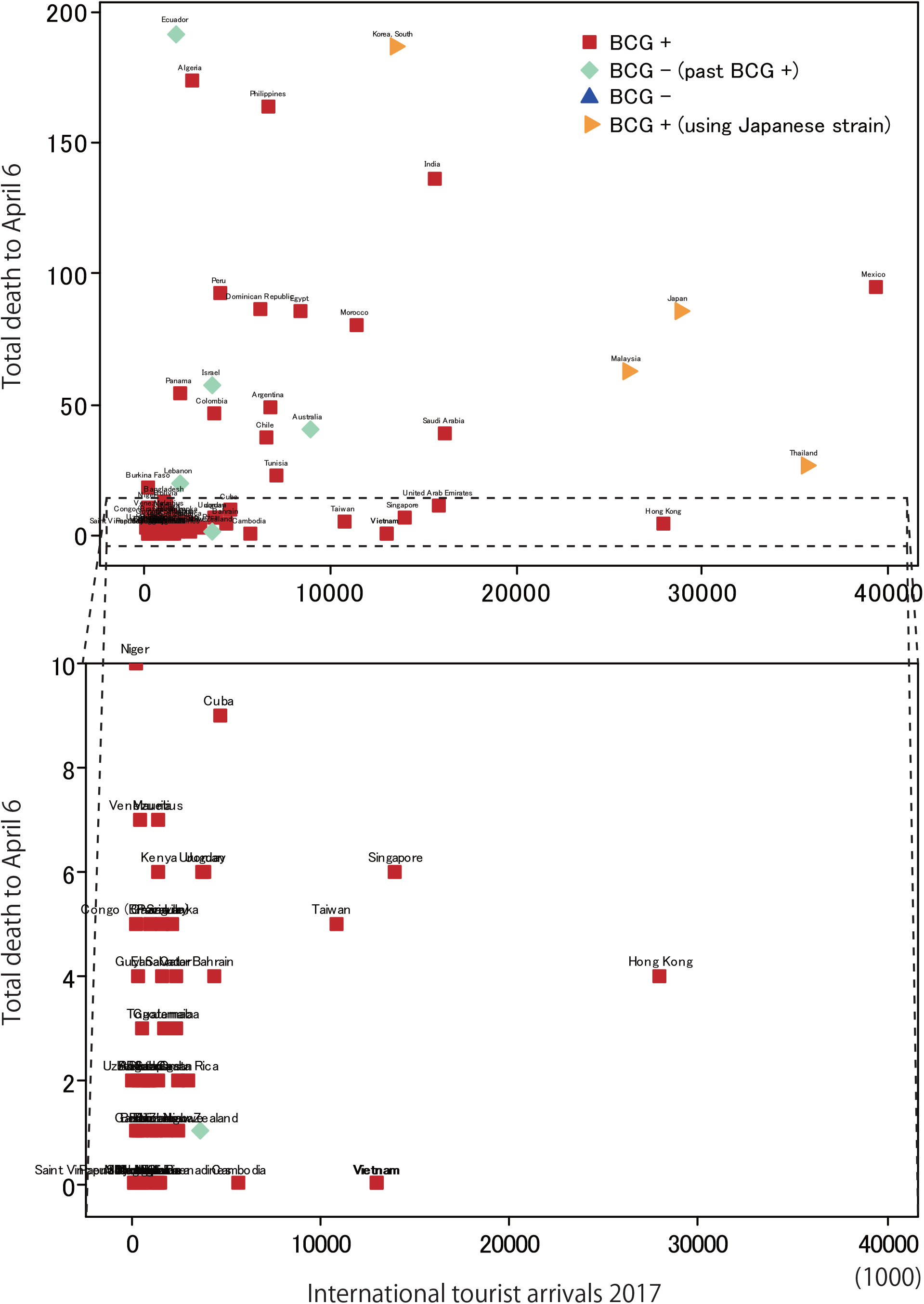
Bivariate plots of International tourist arrivals in 2017 vs. Total deaths up to April 6 using non-European nations with total deaths less than 200. Most nations are BCG +, but some BCG - nations seem to succeed in suppressing COVID-19. Most non-European nations are BCG + and are located where COVID-19 has not yet entered at full scale. This fact would flaw the results of previous studies.

The timing of disease entry seems to correlate to number of tourists (Fig. 3). In addition, political and economic relationships to China might affect disease entry such as in Iran and South Korea. Therefore, the international flow of people might affect the timing of pandemic entry and the number of deaths. However, focusing on nations whose deaths are relatively small, there seems to be less relationship between the number of tourists and total deaths (Fig. 5). Some nations using the Japanese BCG strain such as Thailand, seemed to succeed in preventing transmission despite the number of tourists. However, some BCG- nations, such as Australia and New Zealand, also seem successful in preventing the pandemic. In addition, most nations where the disease spread has so far been prevented are BCG+ nations. These nations are located outside of Europe, which might flaw previous studies (Fig. 5).

Although previous reports suggested that past BCG vaccination reduces infection and mortality (e.g. Miller et al. 2020; Sala and Miyakawa 2020), the present study suggests that these previous results were biased by the route of pandemic spread and the timing of pandemic entry to the nation. In other words, the fact that some nations in Western Europe became an epicenter of the pandemic as a stochastic event is an important factor. This is clearer in the analysis using only European nations (Table 4; Fig. 4).

The present study did not consider GDP, latitude, or temperature, etc. However, the results indicate that even if past BCG vaccination may have an effect on COVID-19, it can be easily masked by other factors. In fact, the timing of pandemic entry had much larger effect on the observed cross-national difference than the BCG policy did (Table 3 and 4). Therefore, the author considers that the effect of BCG, if any, may not be potent as previous reports suggested; the result does not support that the past BCG vaccination reduces the number of infection and death changes by double or triple digits (e.g. Miller et al. 2020; Sala and Miyakawa 2020).

### BCG may possess little or no effect on the increase rate of deaths in countries where the disease spreading

The result of the increase rate of deaths is shown in Fig. 6. The real increase graph is shown in Fig. 7. There is no significant difference between BCG+ or – nations (Fig. 6). However, when analyzing the Japanese BCG strain separately, the result was weakly significant (Fig. 6). The result may be affected by sample size (the comparison was performed on 55 nations). The increase rate decreases from BCG –, BCG – (past +), BCG +, and BCG + (Japanese strain) nations. However, there are several outliers and large regional differences. In addition, few nations affected the result. Furthermore, the timing of the pandemic entry may affect because people’s behavior may have gradually changed. Therefore, the effect of BCG vaccination is not clear. It should be noted that another study reported a highly significant correlation between BCG vaccination and the increasing rate (Akiyama and Ishida 2020). This difference may be attributed to the previous study focusing on the average increase rate whereas the present study is focused on the maximum increase rate. The author considers that the maximum increase rate indicates potential of disease increase and may be more critical to pandemic control.

**Fig. 6.**
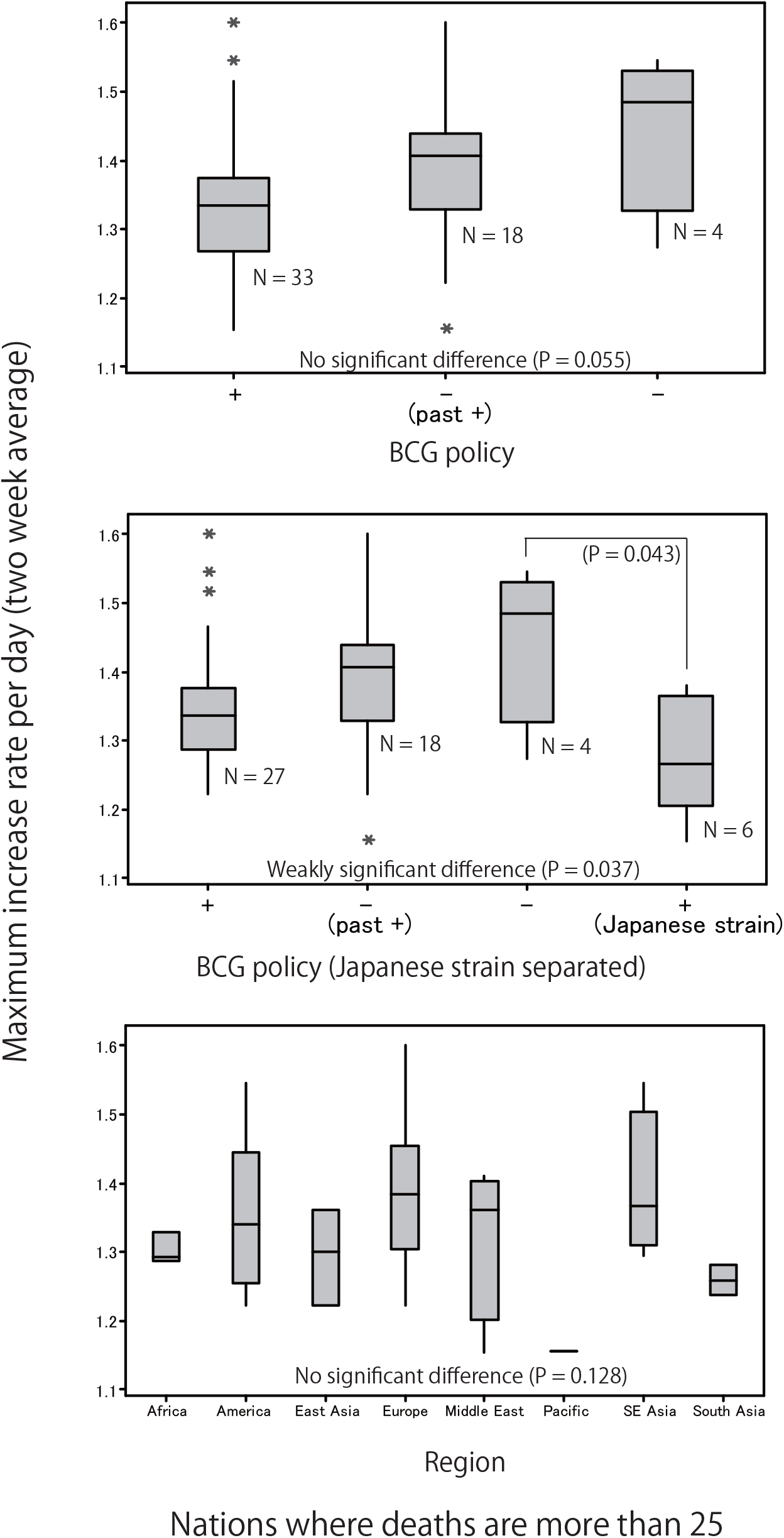
Box plots of maximum increase rate per day (two week average). Nations where deaths are more than 25 were illustrated. Boxes indicate quartiles, central-lateral bars indicate medians, vertical bars indicate the range of the specimens, and asterisks indicate outliers. The increase rate seems to higher in BCG – nations. However, only one pair was weakly statistically significant, but most are not (ANOVA/ Tukey’s test).

**Fig. 7.**
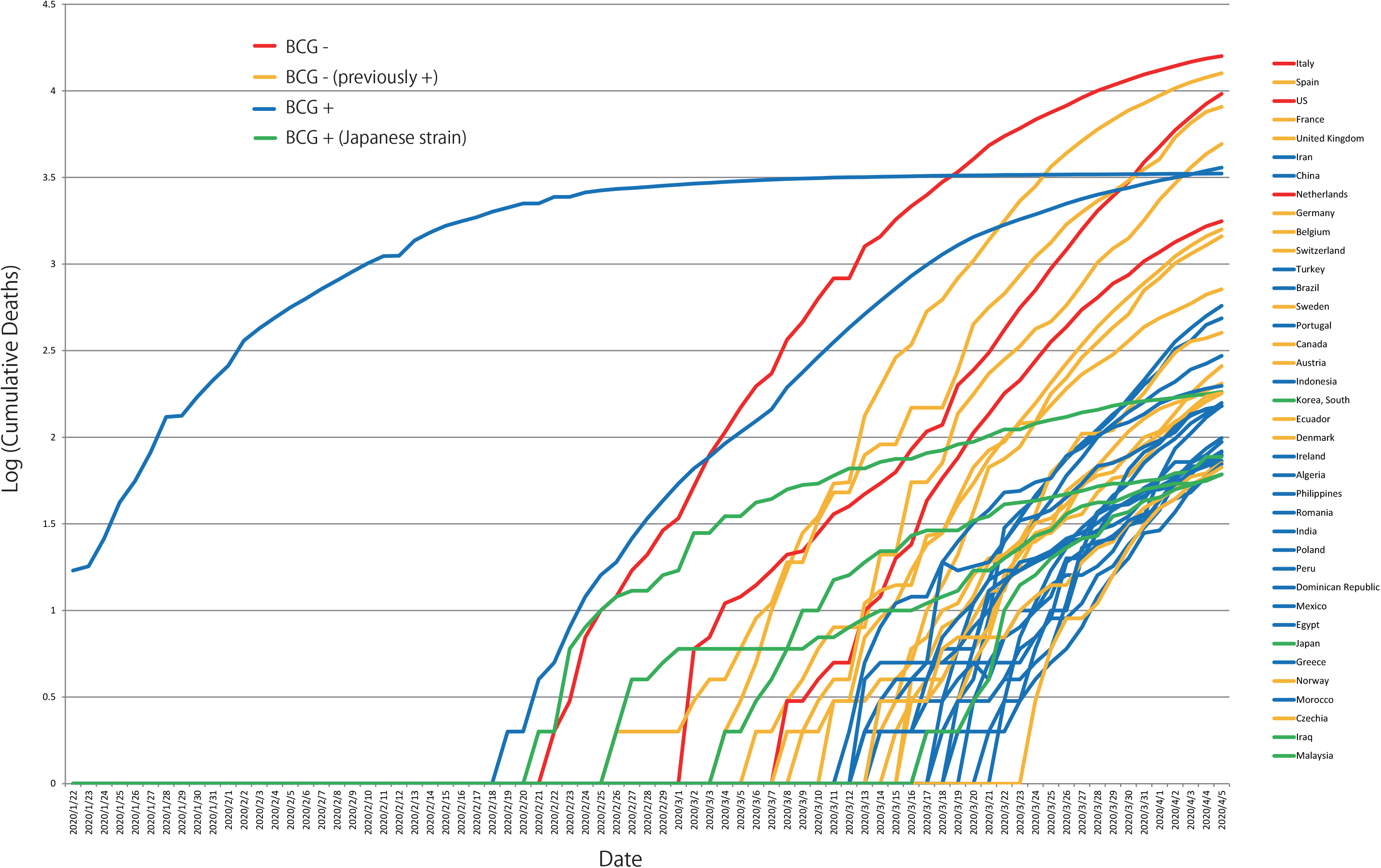
Cumulative deaths of nations in which total deaths over 25. Although the increase rate in Iraq, Korea, and Japan seem to low, at the beginning of the increase, the rate is not particularly low. Malaysia which uses Japanese strain shows a similar pattern to the other nations.

### Possible factors affecting the results and the remaining possibility of the BCG hypothesis

The hypothesis that past BCG vaccination affects COVID-19 can be separated into three. The first hypothesis is regarding reducing personal risk of infection or mortality. The present study partially tested this using the data of 1) the Diamond Princess and did not show positive result. Another study also reported a negative result by comparing infection rate of BCG+ and BCG- age people in several nations (Fukui et al. 2020). The second hypothesis is regarding reducing the total morbidity and mortality in the population (e.g. Miller et al. 2020; Sala and Miyakawa 2020). The present study did not show a positive perspective regarding this aspect, as seen by the result of 2) the cross-national comparisons. The third hypothesis is about reducing the rate of propagation in the population (e.g. Akiyama and Ishida 2020). The present study did not show clear results about this. As discussed above, although the possibility of undetectable effect cannot be ruled out, none of the hypothetical effects of past BCG vaccination on COVID-19 are supported in this study.

The hypotheses of BCG have emerged from previous reports that BCG has non-specific provocation of natural immunity (termed “trained immunity”; reviewed by e.g. Covián et al. 2019; Angelidou et al. 2020). This mechanism is very attractive, although the present study did not show a positive result. However, if this mechanism is effective, then the effects of BCG on COVID-19 may not differ in high magnitude among BCG strains.

T-cell epitopes of BCG have been estimated previously (Zhang et al. 2013). According to the previous study (Zhang et al. 2013), the Japanese BCG strain possesses more epitopes than other strains. The author preliminary examined whether SARS-CoV-2 possesses homologous amino acid sequences of these BCG epitopes using BLAST. However, no homologous sequence was observed (Personal Observations; Fig. 8). If the Japanese strain had a larger effect on the disease, it would not be due to affinity to T-cell receptors. A possible mechanism could be that a domain or constituent unique to the Japanese strain has a high affinity to Toll-like receptors or some other receptors for innate immunity and leads to strong trained immunity (Fig. 8). However, according to the results of the Diamond Princess, this is rather unlikely.

**Fig. 8.**
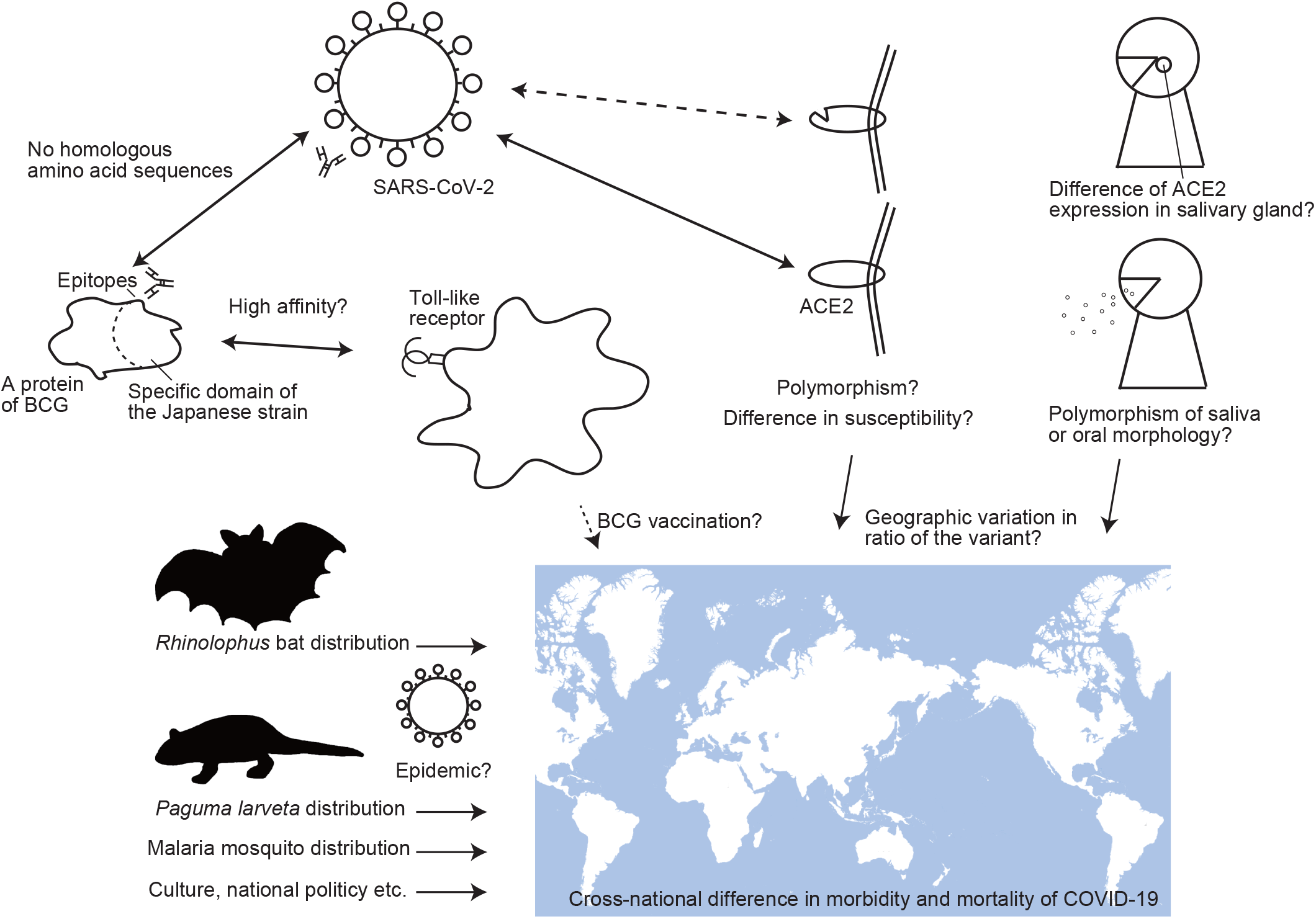
Schematic illustration of how cross-national statistical analysis (“ecological study”) can be influenced. Many factors influence the results. The author’s idea of the remaining possibility of BCG vaccination is also illustrated. The map is modified based on the map provided by the Geospatial Information Authority of Japan. https://maps.gsi.go.jp/development/ichiran.html

The cross-national difference of morbidity and mortality can be influenced by many factors. A schematic illustration is shown in Fig. 8. A possibility explained above is that several nations have unique reasons that reduce disease propagation. Each nation’s policy in response to the pandemic, cultural differences, or lifestyle differences should be considered. Additionally, as our cross-national comparisons showed the region significantly affected the result (Table 3), there can be other hypotheses.

The author could suggest another possible hypothesis that several nations already experienced local epidemics of similar viruses. Wild bats and civets possess viruses related to SARS-CoV-2 (e.g., bat/ civet SARS coronaviruses) (e.g. Guan et al. 2003; Lau et al. 2005; Li et al. 2005; Song et al. 2005; Wu et al. 2016; Luk et al. 2019; Lam et al. 2020). Epitopes of SARS-CoV-2 were suggested by a previous study (Ahmed et al. 2020). The author preliminarily examined whether SARS-related viruses possess homologous amino acid sequences of these SARS-CoV-2 epitopes using BLAST. There are many homologous sequences in bat and civet SARS coronaviruses (Personal Observations). *Rhinolophus* bats are one of the wild hosts of these viruses and distribute in South Europe, Middle East, South Asia, South East Asia, East Asia, and Oceania. Another host is the masked palm civet (*Paguma larvata*) distributes in East Asia. If there were local epidemics of these related viruses manifested as a slight cold in several particular nations, many people may have acquired immunity to these viruses, hence explaining the different morbidity and mortality patterns between nations. A large scale antibody test may address these hypotheses. In addition, cross-immunity to normal colds should also be considered. In Japan, there is a culture that people go to work even if they have a cold. Therefore, a high percentage of the working generation may have already contracted the common cold. This may have an impact on the situation in Japan. However, this effect will not work for the second wave predicted in the near future, as immunity of normal cold does not last long.

The author could suggest one more possible hypothesis being that human genetic variation caused the difference between nations. As other SARS-like coronaviruses, SARS-CoV-2 may use the angiotensin-converting enzyme II (ACE2) protein (Ge et al., 2013; Hoffman et al. 2020) and the transmembrane serine protease (TMPRSS2) protein (Matsuyama et al. 2010; Glowacka et al. 2011; Shulla et al. 2011) for infection. Some reports have suggested that *ACE2* variants (Elisa et al. 2020) or *TMPRSS2* variants (Asselta et al. 2020) can affect the severity of COVID-19. A previous report suggested that the proportion of *ACE2* polymorphisms is different among populations; the ratio of some allele variant gradually decreases from Europe > America > Africa, South Asia> East Asia (including China) (Cao et al. 2020). Geographic variation of MHC class I (HLA) gene whose susceptibility the COVID-19 may be different was also reported (Nguyen et al. 2020). These patterns may explain the relatively low mortality and increase rate in East Asian nations. Perhaps this geographic pattern may have been caused through natural selection by a possible historical epidemic of SARS related viruses in East Asia or other regions. Given that historical contact with livestock may have been important to resistance to some kind of infectious disease (Diamond 1997), increased resistance to another kind of zoonosis can be observed in areas where are close to biodiversity hotspots. Perhaps the resistance could be a cultural or political propensity.

The author could suggest another possible hypothesis of polymorphisms affecting the genetic ability of disease spread. Although many studies focus on susceptibility of the disease, ability of disease transmission to others would be necessary to examine as a factor on the regional difference. In fact, several results suggest relatively low difference of susceptibility among human populations (Table 1 and 2). Previous studies have suggested that almost all infected persons do not spread COVID-19, but rather only a selected few people spread the disease (Nishiura et al. 2020; Endo 2020). In addition, a previous study reported that viral load in the saliva correlated with age (To et al., 2020), but their result seems to show high variance among individuals. If the ability of disease spread varies among individuals and the proportion of “super-spreaders” is different among nations, the previously reported spread pattern (Nishiura et al. 2020) and the different increase rates of victims among nations can be explained. The author suggests two patterns of hypothetical causation. The first is that the degree of expression of ACE2 or other related genes in the salivary gland or intraoral organs varies. If the expression is high, the number of viruses increases in the saliva. COVID-19 may infect through droplets (Lai et al. 2020) possibly including those blown out during conversation. Therefore, this hypothesis is feasible. In addition, the amount or viscidity of saliva, and morphological character of oral organs might affect disease spread. However, to prove this hypothesis, an intensive examination of the route of infection and disease spread would be needed.

As discussed above, there can be many hypotheses explaining the differences of COVID-19 morbidity, mortality, and how those increase among nations. Given that the effect of the region was significant in analysis 2, simple comparisons among nations may fail to address these problems/hypotheses.

Although there are many things to do before proving the BCG hypothesis, many mass media outlets are reporting the hypothesis, causing people to gradually consider the hypothesis to be true (especially in BCG+ nations such as Japan). Some people even suggested that no special protective measure is needed in Japan. The author seriously worries about this problem because the change in people’s behavior is the most important avenue to prevent viral spread (Flaxman et al. 2020; Ministry of Health, Labour and Welfare, Japan, 2020a; Zhang et al. 2020). Optimism caused by the spread of this hypothesis is a serious problem. We should be cautious that in Japan, even though almost all citizens received BCG vaccination (using the Japanese strain, which may be most effective), cases and deaths are constantly increasing at the beginning of April, 2020.

### The effect of past and recent vaccination should not be confused

The present study focuses on past vaccination (such as that in many decades ago) as in the previous cross-national comparative studies. In the meantime, clinical studies examining the effects of recent vaccination to adults are currently being conducted (e.g. reported by Vrieze, 2020). However, it is dangerous to confuse the effects of recent vaccination with the past vaccination (Asahara, submitted). Even if the cross-national comparison results were positive, it cannot prove the effect of recent vaccination because inoculation to infants may be necessary for effectiveness (e.g. low efficacy of adult inoculation for tuberculosis resistance: Mangtani et al. 2014). Of course, the cross-national comparisons have too many factors to consider, and that alone is not sufficient to prove the BCG effect. On the other hand, even if the results of a clinical study are positive, it cannot prove that the effects will last for decades. In other words, the result of the clinical trials cannot prove that past BCG vaccination is responsible for the cross-national differences in cases and deaths. Other studies would be required such as examining the previous vaccination history of infected people in a country where vaccination is optional. However, clinical studies would provide more reliable data on the effect of BCG vaccination on the COVID-19 than cross-national comparisons.

## Conclusions

The hypothesis that BCG vaccination reduces the infection and mortality of COVID-19 is attractive. However, previous international comparative reports may not prove the hypothesis because many other possibilities can explain the observed pattern. In addition, the present study did not show a positive result. Clinical researches are currently being conducted (e.g. reported by Vrieze, 2020). These ongoing clinical studies should provide a better understanding of this hypothesis. Until then, we should be careful about patterns observed in the statistical data. Moreover, spreading the optimistic view triggered by the hypothesis is rather harmful.

## Data Availability

The present study is based on the public data. Dataset is shown in the supplementary file.

## Acknowledgement

The author thanks anonymous readers who commented on the preprint version of the manuscript (published in April 22, 2020). English of the some part of the manuscript have been corrected by courtesy of Editage service.

